# Clinical manifestations along with biochemical and psychological outcomes of COVID-19 cases in diabetic individuals in Bangladesh

**DOI:** 10.1101/2020.09.24.20200790

**Authors:** Farhana Akter, Adnan Mannan, H. M. Hamidullah Mehedi, Abdur Rob, Shakeel Ahmed, Asma Salauddin, Md. Shakhawat Hossain, Md Mahbub Hasan

## Abstract

**Background and aims:** This study investigated the clinical manifestations, outcomes and long-term complications of COVID-19 inpatients in Bangladesh while emphasizing on individuals having diabetes.

**Methods:** A cross-sectional study was conducted for a sample of COVID-19 inpatients across four different hospitals of Bangladesh between April 1st and June 30, 2020. Variation in clinical characteristics, contact history, comorbidities, treatment pattern, and long-term complications were investigated.

**Results:** There were 734 COVID-19 presentations in this study of which 19.8% of patients had diabetes. 76% were male and 85% of the patients had been administered with all vaccine doses during childhood. The most frequently occurring blood groups among patients with diabetes were B (+) ve (35%) and O (+) ve (31%). Among biochemical parameters, glucose, D-dimer, C-reactive protein (CRP) and Troponin levels were significantly elevated amidst the cohort with diabetes. The frequency of insulin dependent individuals increased three-fold during COVID-19. A number of COVID-19 patients with diabetes have been suffering from long term complications post recovery including pain, discomfort, memory loss and sleep disturbance.

**Conclusion:** Individuals with diabetes have experienced severe manifestation of COVID-19 and post disease complications. Further in-depth studies focused on larger sample sizes are entailed to assess the relationships elaborately.

## 1.0 Introduction

COVID-19 is an infectious, contagious disease of the respiratory system caused by a varying, novel strain of the Severe acute respiratory syndrome-related coronavirus (SARS-CoV) known as the Severe acute respiratory syndrome-related coronavirus 2 (SARS-CoV-2) [1]. Since the inception of its outbreak in Wuhan, China at the end of December 2019, COVID-19 increased its host span in a rapid progression globally and therefore was declared a pandemic by the WHO on March 11, 2020 [1, 2].

SARS-CoV-2 is an enveloped, beta coronavirus consisting of a single stranded positive sense RNA (23-32kb) as its core genetic material [3]. SARS-CoV-2 generally spreads through Human-to-human transmission (HHT) by means of respiratory droplets through direct contact or fomite and aerosols [4, 5]. COVID-19 has a diversified pattern in terms of symptoms, recovery rate and mortality rates across the globe. Predominant manifestations of mild flu-like symptoms, shortness of breath, depleted sense of taste and smell were found in the majority of COVID-19 patients [6, 7]; however in some, there were no symptoms at all while in others, the disease either progressed towards more severe clinical complexities including Pneumonia [8, 9], Acute Respiratory Distress Syndrome (ARDS), multi-organ failure or became lethal [2]. The prospect of risk factor in the case of COVID-19 with regards to age nevertheless is still vague since the disease has been diagnosed in people in almost every age group.

Case-control studies conducted on COVID-19 during the pandemic concluded that comorbidity (persistence of medical conditions including obesity, hypertension and diabetes mellitus) could be a determinant on the progression of COVID-19 in individuals [10]. Although the data is quite limited, recent studies have put forward that the prevalence of diabetes mellitus (DM) and elevated blood glucose levels can act as independent factors of mortality and morbidity related to COVID-19; firstly because individuals with diabetes have a prolonged recovery duration from viral diseases due to having a compromised immune system and secondly for the virus’s ability to sustain itself in an environment with high glucose levels putting individuals with diabetes mellitus at a vulnerable position from the aspect of casualties due to COVID-19 [11-13]. Additionally, some recent studies proclaimed a number of COVID-19 associated long term complications [14, 15] which require further investigation and assessment to establish facts in detail.

With a population of more than 161 million people, Bangladesh stands eighth among the most populated countries in the world [16]. In Bangladesh, as of 24 September, 2020, infections from SARS-CoV-2 has spiked to nearly 352,287 individuals while the death count figured to 5,044 people (https://iedcr.gov.bd/). Among other conventional diseases and disorders in Bangladesh, cases of diabetes are on the rise at an alarming rate and as available data from the International Diabetes Federation (IDF) tell us, there are 8.4 million cases of diabetes in adults in Bangladesh [17]. To date, multiple scientific studies have been conducted and published relevant to the clinical characterization of COVID-19. Be that as it may, the association of COVID-19 with diabetes and the disease’s post recovery effect on individuals is still a subject that needs attention as well as analysis. The first of its kind in Bangladesh from aspects of clinical epidemiology, metabolic changes and long-term effects of COVID-19 on diabetic cases, the aim of this study was to identify the frequency of diabetic individuals within a cohort of COVID-19 diagnosed individuals being provided with medical attention at different health care facilities and to compare the clinical manifestations and long-term complications among the diabetic and non-diabetic COVID-19 patients attending those healthcare facilities.

## 2.0 Methods

### 2.1 Study Design

A cross sectional observational study was conducted among COVID-19 cases confirmed positive by RT-PCR assay seeking medical attention in four different medical institutes. In order to study complications post COVID-19 in individuals, patients who tested negative after two consequent RT-PCR assessments at an interval of 24 hours and 4 weeks prior to the interview date were considered.

### 2.2 Study Sites

The study was conducted throughout four hospitals providing aid to individuals diagnosed with COVID-19. Located in the Chattogram division of Bangladesh, the institutes were: Chittagong General Hospital, Chittagong Medical College Hospital, Bangladesh Institute of Tropical and Infectious Diseases (BITID) and Chattogram Field Hospital. The tenure of the study was from April 2020 to June 2020.

### 2.3 Case Definition

The presence or absence of SARS-CoV-2 infection and by extension, a positive or negative case of COVID-19 was confirmed by RT-PCR assay. Individuals who resided in the hospitals for more than 24 hours were considered to be inpatients. Cases having a glycated haemoglobin (HbA1c) content of 6.5% who went through any approved biochemical assessment of diabetes mellitus previously were considered within the group having diabetes. Two or more blood glucose level assessments which returned a result greater than 11.1 mmol/l defined uncontrolled hyperglycemia; contrary to which blood glucose levels < 4 mmol/l classified as hypoglycaemia. Samples were split depending on them having or not having diabetes considering medication itinerary of antidiabetic drugs or fasting blood glucose level during admittance in the hospital as determinants.

### 2.4 Sample size & Data collection

A total of 734 patients diagnosed with COVID-19 were interviewed for this study. A relevant questionnaire and medical records were taken into account as the principle sources of data. In terms of data, hospital records were also reviewed and history of vaccination was collected through self-reporting as well as records. All retrospective data collected by physicians over telephone interviews were manually put into a Google Form. All information added on the form which represented answers given by the subjects were double checked, submitted and the records were preserved. The questionnaire in use asked for a patient’s socio-demographic information, clinical manifestations, biochemical parameters, behavioral practices, comorbidities, medication types, laboratory tests, electrocardiogram results, inpatient medications, treatments and outcomes (including length of stay in the hospital, discharge, re-admission, and mortality).

### 2.7 Exclusion Criteria

Deceased patients and those who had no interest in participation or had concerns and disagreed to provide consent were excluded.

### 2.8 Ethical Consideration

Preceding the interview, verbal consent was taken from the subjects while a written consent waiver was received from Institutional Review Board (IRB) for this study. The protocol was then approved by IRB of Chattogram General Hospital. The ethical approval number for this study is 2020-03-027.

### 2.9 Statistical Analysis

Descriptive and inferential statistics were used to analyze the collected data. In order to present categorical variables frequency counts and percentages were used. Association between categorical variables were evaluated using Pearson’s chi-square test with continuity correction according to necessity. All analyses were conducted using SPSS Statistics 25 (IBM, Armonk, New York) licensed to King’s College London, UK.

## 3.0 Results

### 3.1 Demography and socio-economic conditions of COVID-19 patients with and without Diabetes

A total of 734 cases were reported in this study of which, 80.11% were without diabetes and 19.89% were with diabetes. Manifestations of COVID-19 among individuals with diabetes and without diabetes significantly varied among different age groups (p= 0.000) (Table-1). Although one-third of the COVID-19 patients with diabetes belonged to the elder age group (≥ 60 years), the persistence of COVID-19 was higher in the patients not exceeding 39 years of age and without diabetes (Table-1). The number of male patients (76%) was three times higher than female patients (24%) regardless of them being with or without diabetes. In the dataset, the most frequently occurring blood groups among patients with diabetes were B (+) ve (35%) and O (+) ve (31%), while there were no A (-) ve and B (-) ve blood groups among the individuals. The prevalence of COVID-19 cases was found to be comparatively higher in the urban area (70%). Monthly gross family income and burden of diabetes among COVID-19 patients were significantly correlated (p=0.000). We found that about 40% COVID-19 patients belonged to middle-income families with a gross monthly income between BDT 25,000 and BDT 49,999 (296-590 USD). Interesting enough, COVID-19 patients from the low-income tier had a lower burden of diabetes (18.5% vs 30.4%) whereas the high-income group had almost double the burden of diabetes (23.3% vs 11.2%). There were no significant differences in terms of being vaccinated or not vaccinated among the patients with and without diabetes. The majority of COVID-19 cases were found to have been administered with all vaccines (91%) and BCG vaccine (85%) but this does not have any impact on the outcome of the disease depending on them having diabetes or not having diabetes. The most common comorbidities were cardiovascular diseases (24%), respiratory diseases (11%), and history of heart attack (10%) in patients with diabetes. Moreover the frequency of comorbidities, cancer (3.4% vs 0.9%, p=0.016), cardiovascular disease (24% vs 5.4%, p=0.000), respiratory disease (11% vs 5%, p= 0.007), liver disease (5.5% vs 1%, p=0.002), history of heart attack (10% vs 0.71%, p=0.000) and other chronic disease (14% vs 6.5%, p=0.004) are statistically significant in COVID-19 patients with diabetes (Table-1).

**Table 1:** Characteristics of study subjects for all patients and patients with and without diabetes.

| **Variables** | **Without Diabetes** | **With Diabetes** | **Total** | **Pearson's $\chi^2$** | **P value** |
| --- | --- | --- | --- | --- | --- |
|  | 588 | 146 | 734 |  |  |
| ***Age (years)*** |  |  |  |  |  |
| 0-9 | 15 (2.6%) | 0 (0.0%) | 15 (2.0%) | 171.723 | 0.000 |
| 10-19 | 35 (6.0%) | 0 (0.0%) | 35 (4.8%) |  |  |
| 20-29 | 169 (28.7%) | 4 (2.7%) | 173 (23.6%) |  |  |
| 30-39 | 193 (32.8%) | 17 (11.6%) | 210 (28.6%) |  |  |
| 40-49 | 86 (14.6%) | 42 (28.8%) | 128 (17.4%) |  |  |
| 50-59 | 58 (9.9%) | 39 (26.7%) | 97 (13.2%) |  |  |
| ≥60 | 32 (5.4%) | 44 (30.1%) | 76 (10.4%) |  |  |
| ***Sex*** |  |  |  |  |  |
| Female | 142 (24.1%) | 34 (23.3%) | 176 (24.0%) | 0.048 | 0.827 |
| Male | 446 (75.9%) | 112 (76.7%) | 558 (76.0%) |  |  |
| ***Blood group*** |  |  |  |  |  |
| A (-) ve | 9 (1.5%) | 0 (0.0%) | 9 (1.2%) | 18.130 | 0.011 |
| A (+) ve | 121 (20.6%) | 32 (21.9%) | 153 (20.8%) |  |  |
| AB (-) ve | 4 (0.7%) | 2 (1.4%) | 6 (0.8%) |  |  |
| AB (+) ve | 57 (9.7%) | 11 (7.5%) | 68 (9.3%) |  |  |
| B (-) ve | 10 (1.7%) | 0 (0.0%) | 10 (1.4%) |  |  |
| B (+) ve | 221 (37.6%) | 51 (34.9%) | 272 (37.1%) |  |  |
| O (-) ve | 1 (0.2%) | 4 (2.7%) | 5 (0.7%) |  |  |
| O (+) ve | 165 (28.1%) | 46 (31.5%) | 211 (28.7%) |  |  |
| ***Dwelling type*** |  |  |  |  |  |
| Urban | 407 (69.2%) | 108 (74.0%) | 515 (70.2%) | 1.263 | 0.261 |
| Village | 181 (30.8%) | 38 (26.0%) | 219 (29.8%) |  |  |
| ***Monthly income (BDT)*** |  |  |  |  |  |
| ≤9,999 | 42 (7.10%) | 7 (4.80%) | 49 (6.70%) | 21.268 | 0.000 |
| 10,000-24,999 | 179 (30.40%) | 27 (18.50%) | 206 (28.10%) |  |  |
| 25,000-49,999 | 237 (40.30%) | 56 (38.40%) | 293 (39.90%) |  |  |
| 50,000-74,999 | 64 (10.90%) | 22 (15.10%) | 86 (11.70%) |  |  |
| ≥75k | 66 (11.20%) | 34 (23.30%) | 100 (13.60%) |  |  |
| ***BCG vaccine*** |  |  |  |  |  |
| Yes | 509 (86.6%) | 119 (81.5%) | 628 (85.6%) | 2.421 | 0.12 |
| No | 79 (13.4%) | 27 (18.5%) | 106 (14.4%) |  |  |
| ***All Vaccination*** |  |  |  |  |  |
| Yes | 542 (92.2%) | 128 (87.7%) | 670 (91.3%) | 2.983 | 0.084 |
| No | 46 (7.8%) | 18 (12.3%) | 64 (8.7%) |  |  |
| **History of contacts with COVID-19 patients** |  |  |  |  |  |
| Direct contact with COVID-19 patients | 309 (52.6%) | 55 (37.7%) | 364 (49.6%) | 10.359 | 0.001 |
| Indirect contact with COVID-19 patients | 345 (58.7%) | 76 (52.1%) | 421 (57.4%) | 2.095 | 0.148 |
| Contact with foreign returnees | 31 (5.3%) | 8 (5.5%) | 39 (5.3%) | 0.01 | 0.92 |
| Go outside of home | 432 (73.5%) | 93 (63.7%) | 525 (71.5%) | 5.482 | 0.019 |
| Had infected family member(s) | 233 (39.6%) | 58 (39.7%) | 291 (39.6%) | 0.000 | 0.982 |

\*BDT= Bangladeshi currency, 1 USD= 84.90 Bangladeshi Taka (as of 08.09.202)

We tried to trace their if they were in close proximity with any confirmed COVID-19 patients directly, indirectly or both. For the record, we even tried to trace their frequency of going out prior infection and whether they were in any proximity with returnees from abroad. We found that 49.6% (n=264) COVID-19 patients had direct contact, 57.4% (n=421) had indirect contact with COVID-19 patients before being infected and 39 (5.3%) had contact with foreign returnees (Table-1). 525 (71.5%) COVID-19 patients went outside of their homes at least once a day and 291 (39.5%) COVID-19 patients had family members who were already infected.

Compared to COVID-19 patients without diabetes, direct contact with any other COVID-19 patients (with diabetes 55, 37.7% vs without diabetes 309, 52.6%, P= 0.001) and going outside their homes (with diabetes 93, 63.7% vs without diabetes 432, 73.5%, P=0.019) were significantly higher in COVID-19 patients with diabetes. Patients without diabetes went out (at least once a day) more significantly (P <0.05) than patients with diabetes prior to getting infected. In addition to that patients without diabetes had direct contact with COVID-19 positive patients (at least once) more significantly than patients with diabetes. No significant differences were observed between COVID-19 patients with and without diabetes regarding indirect contact with other COVID-19 patients (With diabetes n=76, 52.1% vs without diabetes 345, 58.7%, P=0.148), contact with people returned from foreign (with diabetes n=8, 5.5% vs without diabetes n=31, 5.5%, P=0.92) and SARS-CoV-2 infected family member (with diabetes n=58, 39.7% vs without diabetes n=233, 39.6%, P=0.982).

In the cohort with diabetes, 41% of the cases have a family history of diabetes (Supplementary file-1, Table S1). 25% of them being maternal while the rest of the 16% being paternal. Furthermore, 48% of the COVID-19 cases with diabetes were diagnosed with diabetes more than 5 years ago whereas 19.8% of the patients have been diagnosed less than a year back. Among the subjects, 1.35% patients (n=10) had no previous history of diabetes and were diagnosed with diabetes after being diagnosed with COVID-19 (Supplementary file-1, Table S1).

### 3.2 Variation in clinical manifestations of COVID-19 among patients with and without Diabetes in Bangladesh

Variations in clinical manifestations of COVID-19 patients with and without diabetes have been observed and highlighted in Table 2. 8.9% (n=64) was asymptomatic and among them COVID-19 patients without diabetes (10%, n= 59) were significantly higher (p=0.024) than that of the patients with diabetes (4.1%, n=6).

**Table 2:**
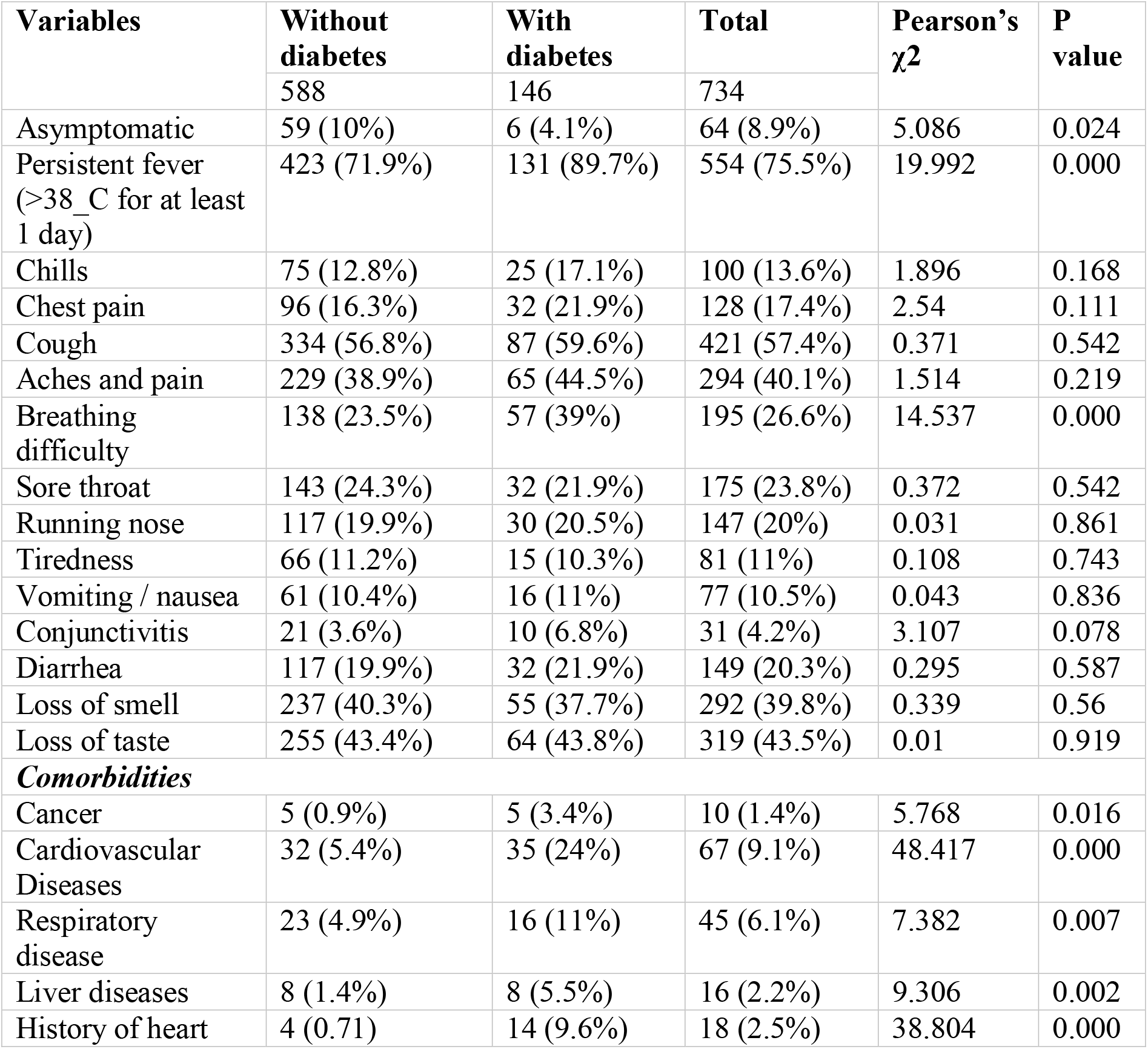
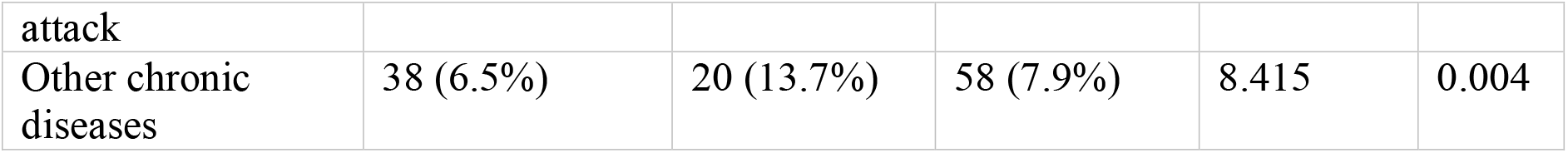
Variation in clinical manifestation of COVID-19 among patients with and without diabetes in Bangladesh (N=734).

The most common symptoms observed on the onset of the disease, regardless of ages and sex, were persistent fever (75.5%, n=554), cough (57.4%, n=421), loss of taste (43.5%, n=319), aches and pain (40.1%, n= 294), loss of smell 39.8% (n=292). Persistent fever (temperature > 38°C) was observed in 75.5% (n=554) patients extending beyond 1 day. It was significantly higher (P < 0.05) in patients with diabetes (89.7%, n=131) than that of the patients without diabetes (71.9%, n=423). Among 195 (26.6%) SARS-CoV-2 positive patients, breathing difficulties including shortness of breath and/or rapid breathing was also significantly higher in COVID-19 patients with diabetes (57, 39%) than that in patients without diabetes (n= 138, 23.5%).

For symptoms, no significant deviations were found when compared between COVID-19 patients with and without diabetes such as chills, chest pain, cough, aches and pain, sore throat, running nose, tiredness, vomiting, conjunctivitis, diarrhea, loss of smell, loss of taste and hair-fall. Individuals within the 30-39 age group had significantly more fever, body ache, cough and breathing difficulties (Supplementary file-1, Figure S1). Symptoms like sore throat, loss of smell and taste were observed more in COVID-19 individuals aged 20-29 (Supplementary file-1, Figure S1).

### 3.3 Biochemical parameters of COVID-19 patients with and without Diabetes

In this study, various biochemical and hematological parameters were scrutinized among COVID-19 patients with diabetes and without diabetes (Table-3). For COVID-19 patients, individuals with diabetes were significantly more hyperglycemic (P<0.05) than without diabetes. In addition to that, serum creatinine (15.9% vs 4.5%), CRP (44% versus 19%), Troponin (43.2% vs 15%) and the Total count of WBC, ESR were significantly (P<0.05) higher in COVID-19 patients with diabetes than those without diabetes. However, the degree of serum uric acid was lower in COVID-19 cases with diabetes. The TC counts of WBC of most of the patients were in the normal range, but the classified count showed that the TC counts of COVID-19 patients with diabetes were significantly higher than those without diabetes. Although the platelet count of most of the patients were within the normal range, the categorical data showed that it was significantly (P <0.05) higher among the COVID-19 cases with diabetes. Compared to cases without diabetes, the level of D-dimer, troponin and CRP was significantly higher in patients with diabetes with no significant differences in the level of Ferritin and SGPT. However, more than 60% of the COVID-19 cases with and without diabetes were found to have more high Ferritin levels.

**Table 3:** Laboratory and radiological findings of COVID-19 patients at admission:

| **Variables** | **Without diabetes** | **With diabetes** | **Total** | **Pearson's $\chi^2$** | **P value** |
| --- | --- | --- | --- | --- | --- |
| *Blood sugar level (n=734)* |  |  |  |  |  |
| < 4 mmol/L | 1 (0.2%) | 2 (1.4%) | 3 (0.4%) | 82.983 | 0.000 |
| 4-11 mmol/L | 496 (84.4%) | 72 (49.3%) | 568 (77.4%) |  |  |
| $\geq 11.1$ mmol/L | 91 (15.5%) | 72 (49.3%) | 163 (22.2%) | | |
| *Serum uric acid (n=229)* |  |  |  |  |  |
| High | 19 (19.6%) | 10 (7.6%) | 29 (12.7%) | 7.294 | 0.007 |
| Normal | 78 (80.4%) | 122 (92.4%) | 200 (87.3%) |  |  |
| *Serum creatinine (n=219)* |  |  |  |  |  |
| High | 5 (4.5%) | 17 (15.9%) | 22 (10%) | 7.902 | 0.005 |
| Normal | 107 (95.5%) | 90 (84.1%) | 197 (90%) |  |  |
| *CRP (n=390)* |  |  |  |  |  |
| High | 52 (19.2%) | 53 (44.2%) | 105 (26.9%) | 32.03 | 0.000 |
| Normal | 218 (80.8%) | 67 (55.8%) | 285 (73.1%) |  |  |
| *Troponin (n=214)* |  |  |  |  |  |
| High | 21 (15%) | 32 (43.2%) | 53 (24.8%) | 20.725 | 0.000 |
| Normal | 119 (85%) | 42 (56.8%) | 161 (75.2%) |  |  |
| *Ferritin (n=361)* |  |  |  |  |  |
| High | 141 (62.9%) | 84 (61.3%) | 225 (62.3%) | 2.075 | 0.354 |
| Low | 3 (1.3%) | 0 (0.0%) | 3 (0.8%) |  |  |
| Normal | 80 (35.7%) | 53 (38.7%) | 133 (36.8%) |  |  |
| *D-Dimer (n=416)* |  |  |  |  |  |
| High | 139 (47.3%) | 79 (64.8%) | 218 (52.4%) | 10.557 | 0.001 |
| Normal | 155 (52.7%) | 43 (35.2%) | 198 (47.6%) |  |  |
| *SGPT (n=290)* |  |  |  |  |  |
| High | 81 (45.3%) | 49 (44.1%) | 130 (44.8%) | 0.034 | 0.854 |
| Normal | 98 (54.7%) | 62 (55.9%) | 160 (55.2%) |  |  |
| *CBC WBC TC (n=203)* |  |  |  |  |  |
| High | 16 (11.8%) | 20 (29.9%) | 36 (17.7%) | 10.425 | 0.005 |
| Low | 1 (0.7%) | 0 (0.0%) | 1 (0.5%) |  |  |
| Normal | 119 (87.5%) | 47 (70.1%) | 166 (81.8%) |  |  |
| *Lymphocyte (n=203)* |  |  |  |  |  |
| High | 0.7% | 3 (4.5%) | 4 (2.0%) | 3.258 | 0.196 |
| Low | 66 (48.5%) | 31 (46.3%) | 97 (47.8%) |  |  |
| Normal | 69 (50.7%) | 33 (49.3%) | 102 (50.2%) |  |  |
| *Neutrophil (n=203)* |  |  |  |  |  |
| High | 70 (51.5%) | 34 (50.7%) | 104 (51.2%) | 0.266 | 0.876 |
| Low | 1 (0.7%) | 1 (1.5%) | 2 (1.0%) |  |  |
| Normal | 65 (47.8%) | 32 (47.8%) | 97 (47.8%) |  |  |
| *Platelet (n=203)* |  |  |  |  |  |
| Low | 7 (5.1%) | 8 (11.9%) | 15 (7.4%) | 3.027 | 0.082 |
| Normal | 129 (94.9%) | 59 (88.1%) | 188 (92.6%) |  |  |
| *ESR (n=213)* |  |  |  |  |  |
| High | 9 (8.2%) | 32 (31.1%) | 41 (19.2%) | 17.924 | 0.000 |
| Normal | 101 (91.8%) | 71 (68.9%) | 172 (80.8%) |  |  |

\*Reference value: D-dimer < 0.5 µg/mL, Ferritin -Male 18-370 ng/mL, Female 9- 120ng/mL; Serum Creatinine- Male 0.4-1.4 mg/dL, Female - 0.3mg -1.1mg/dL; CRF: < 5mg/L; Troponin: < 0.30 ng/mL; Uric acid: Male- 3.4 – 7 mg/dL, Female 2.4 – 6 mg/dL; ESR: Male < 22 mm/hr, Female < 29mm/hr; SGPT: Male: 15- 65 U/L, Female 15 -60 U/L; CBC : WBC: 4-11 x 10<sup>9</sup>/L, Neutrophil: 40-75%; lymphocyte 20-45%, ESR: male 0-10 mm, Female 0-15 mm.

### 3.4 Medication itinerary in COVID-19 cases with and without diabetes

The percentage of cases with diabetes taking insulin as an aid dramatically increased (more than three times) when diagnosed with COVID-19. Prior to being infected, only 18% of the individuals took insulin shots, which increased very promptly to 63% post COVID-19 diagnosis (Figure-1). On the contrary, the percentage of intake of other medications related to diabetes by COVID-19 patients except insulin such as Biguanide and Sulfonylurea decreased from 44% to 29% and from 11% to 4% respectively. In addition, intake of DPP4 inhibitors and other drugs were also decreased from 2% to 1%.

**Figure 1:**
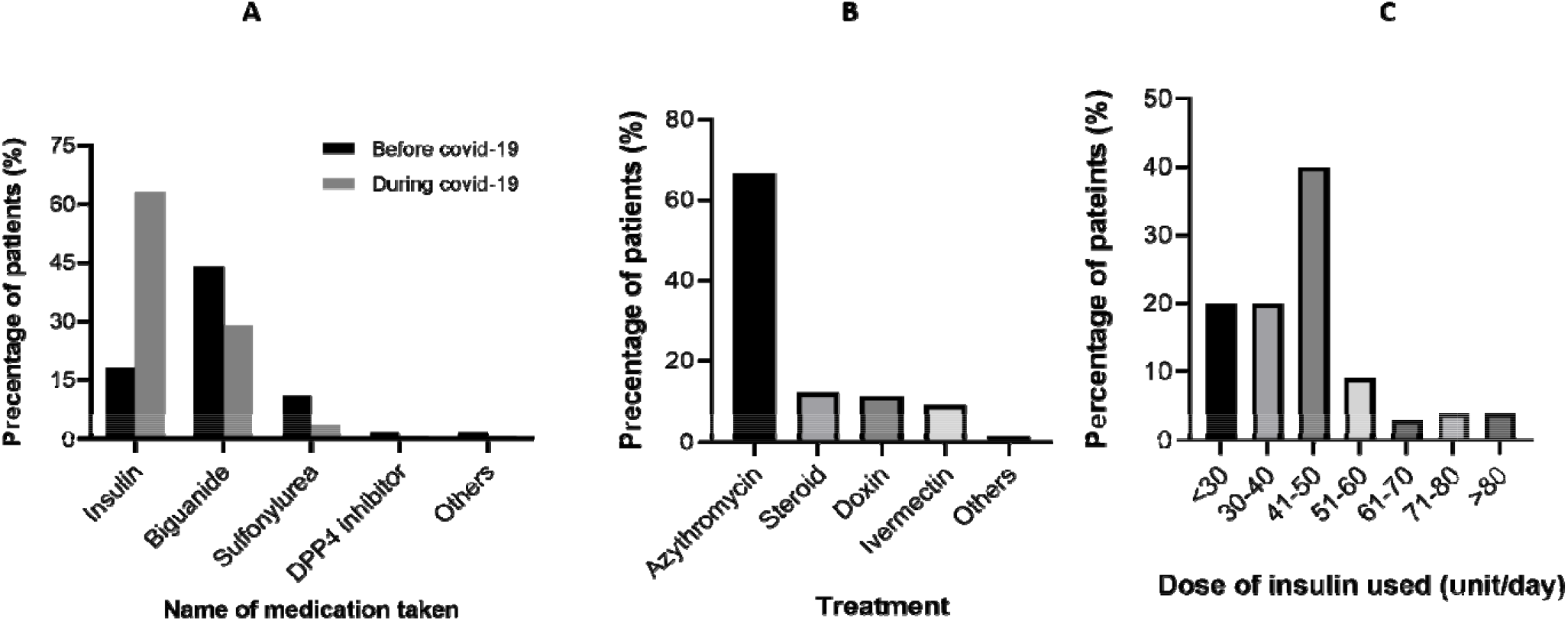
Various treatments and medications of diabetic patients during covid-19. A) Percentage of the anti-diabetic medications used before and during covid-19. B) Frequency of different treatments and drugs during covid-19 C) Dose of insulin used during covid-19.

The increased rate of insulin dependent individuals among COVID-19 groups drew more prudent attention. Graph C shows the insulin doses and percentages of patients in its horizontal and vertical axis respectively. The most frequently taken insulin dose was 41-50 units.

Some common natural remedies along with antibiotics were acclimatized by individuals with COVID-19 regardless of their being diagnosed with diabetes. The antibiotics and medications encompassed Azithromycin (67.0%), Steroids (12%), Doxin (10.4%), Ivermectin (9.0%) and multiple other types (1.6%) (Figure.1).

### 3.5 Physical and mental health complications in recovered COVID-19 cases with and without Diabetes

In this study, the complications of patients after 4 weeks of being recovered from COVID-19 were investigated. Most of the patients (29.8%) were having moderate pain or discomfort (Table-4). However, individuals with diabetes (40%) were found to have significantly (P<0.05) higher levels of pain than those without diabetes (27.3%). Troubles in mobility and movement were significantly (P<0.01) experienced by recovered patients with diabetes than those without. Other complications observed were loss of concentration (24.80%), moderate anxiety or depression (20%), disturbance in sound sleep (19.7%), moderate memory loss (16.7%), panic attacks (13.6%) and hair-fall (8.8%). However no significant differences were observed in terms of the aforesaid complications between groups with and without diabetes.

**Table 4:**
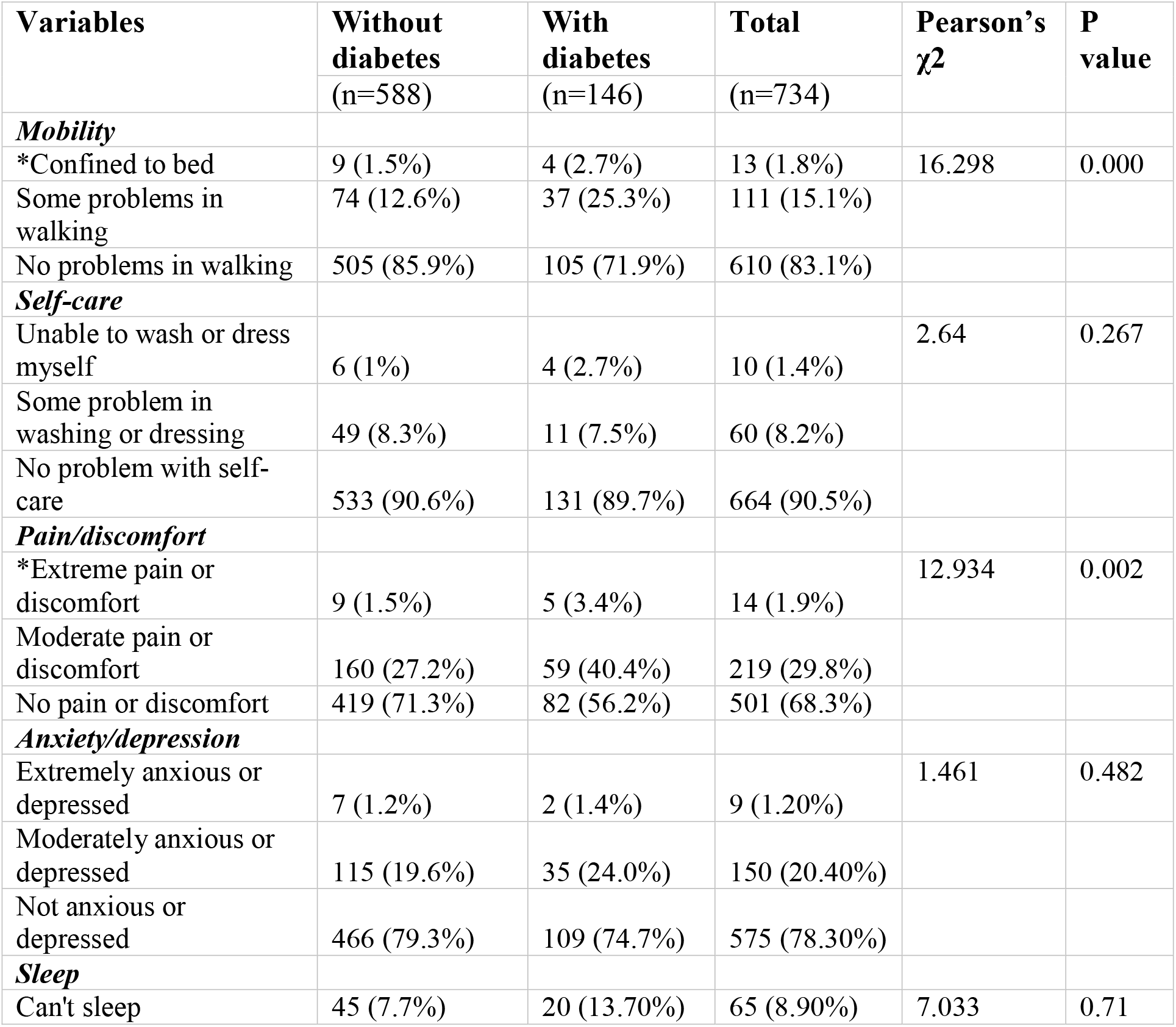
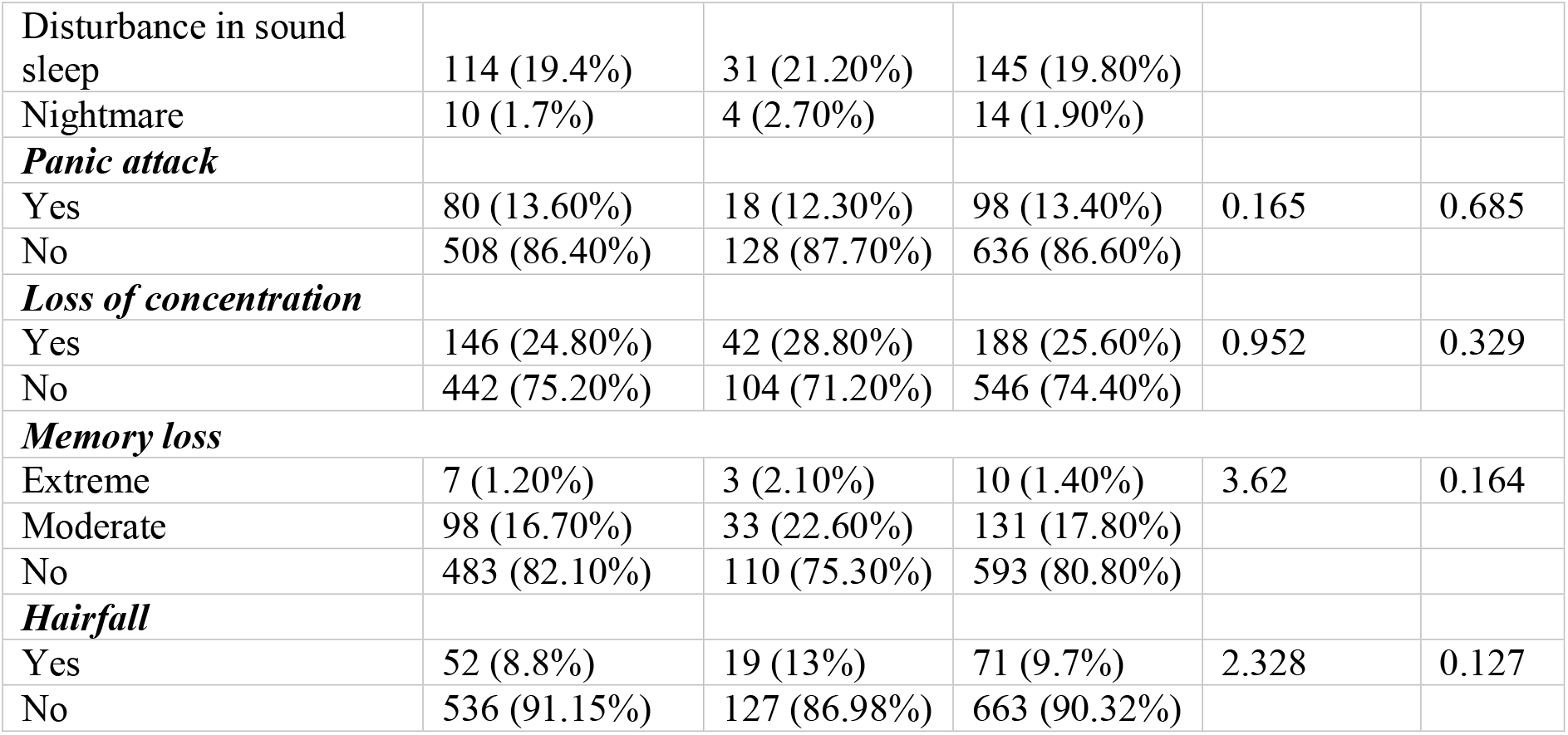
Physical and mental health complications in recovered COVID-19 cases with and without Diabetes:

| **Variables** | **Without diabetes**<br>**(n=588)** | **With diabetes**<br>**(n=146)** | **Total**<br>**(n=734)** | **Pearson's $\chi^2$** | **P value** |
| --- | --- | --- | --- | --- | --- |
| ***Mobility*** |  |  |  |  |  |
| *Confined to bed | 9 (1.5%) | 4 (2.7%) | 13 (1.8%) | 16.298 | 0.000 |
| Some problems in walking | 74 (12.6%) | 37 (25.3%) | 111 (15.1%) |  |  |
| No problems in walking | 505 (85.9%) | 105 (71.9%) | 610 (83.1%) |  |  |
| ***Self-care*** |  |  |  |  |  |
| Unable to wash or dress myself | 6 (1%) | 4 (2.7%) | 10 (1.4%) | 2.64 | 0.267 |
| Some problem in washing or dressing | 49 (8.3%) | 11 (7.5%) | 60 (8.2%) |  |  |
| No problem with self-care | 533 (90.6%) | 131 (89.7%) | 664 (90.5%) |  |  |
| ***Pain/discomfort*** |  |  |  |  |  |
| *Extreme pain or discomfort | 9 (1.5%) | 5 (3.4%) | 14 (1.9%) | 12.934 | 0.002 |
| Moderate pain or discomfort | 160 (27.2%) | 59 (40.4%) | 219 (29.8%) |  |  |
| No pain or discomfort | 419 (71.3%) | 82 (56.2%) | 501 (68.3%) |  |  |
| ***Anxiety/depression*** |  |  |  |  |  |
| Extremely anxious or depressed | 7 (1.2%) | 2 (1.4%) | 9 (1.20%) | 1.461 | 0.482 |
| Moderately anxious or depressed | 115 (19.6%) | 35 (24.0%) | 150 (20.40%) |  |  |
| Not anxious or depressed | 466 (79.3%) | 109 (74.7%) | 575 (78.30%) |  |  |
| ***Sleep*** |  |  |  |  |  |
| Can't sleep | 45 (7.7%) | 20 (13.70%) | 65 (8.90%) | 7.033 | 0.71 |
| Disturbance in sound sleep | 114 (19.4%) | 31 (21.20%) | 145 (19.80%) |  |  |
| Nightmare | 10 (1.7%) | 4 (2.70%) | 14 (1.90%) |  |  |
| **Panic attack** |  |  |  |  |  |
| Yes | 80 (13.60%) | 18 (12.30%) | 98 (13.40%) | 0.165 | 0.685 |
| No | 508 (86.40%) | 128 (87.70%) | 636 (86.60%) |  |  |
| **Loss of concentration** |  |  |  |  |  |
| Yes | 146 (24.80%) | 42 (28.80%) | 188 (25.60%) | 0.952 | 0.329 |
| No | 442 (75.20%) | 104 (71.20%) | 546 (74.40%) |  |  |
| **Memory loss** |  |  |  |  |  |
| Extreme | 7 (1.20%) | 3 (2.10%) | 10 (1.40%) | 3.62 | 0.164 |
| Moderate | 98 (16.70%) | 33 (22.60%) | 131 (17.80%) |  |  |
| No | 483 (82.10%) | 110 (75.30%) | 593 (80.80%) |  |  |
| **Hairfall** |  |  |  |  |  |
| Yes | 52 (8.8%) | 19 (13%) | 71 (9.7%) | 2.328 | 0.127 |
| No | 536 (91.15%) | 127 (86.98%) | 663 (90.32%) |  |  |

## 4.0 Discussion

Characterization of 734 COVID-19 patients was conducted in the current study considering the sample’s demographics, clinical manifestations, preceding comorbidities that were chronic in nature, treatment and interim prognosis. Our study pinpointed that a relatively high prevalence of diabetes (19.89%) was found in individuals diagnosed with COVID-19 and that the prevalence of diabetes altered several biomarkers, exemplified severe clinical subtypes during disease exposition and exhibited a more critical prognosis when compared to COVID-19 patients without diabetes. To the best of our knowledge, this study pioneered in investigating the clinical characteristics and prognosis of COVID-19 patients with diabetes in Bangladesh. For the past few decades, the prevalence of diabetes mellitus in Bangladesh has been on the rise. Although previous studies reported a 9% to 14% prevalence of diabetes in COVID-19 patients [14, 18, 19], our study concluded on a higher prevalence rate for the same. However, a larger proportion of geriatric patients in our sample might contribute to the higher prevalence rate of diabetes that we found in our study. Similar to previous studies in other regions [20, 21] our study also found that 76% of the diagnosed cases were males and that the frequency of COVID-19 was significantly higher in individuals with diabetes aged 40 and higher. Apart from diabetes mellitus, cardiovascular diseases (24%), respiratory diseases (11%) and chronic diseases were also found to be typical chronic illnesses in the sample for this study.

Nuance in expression of blood group antigen can either elevate or reduce host susceptibility to infections caused by Helicobacter pylori, Norwalk virus, and SARS-CoV [22]. By serving as receptors and/or co-receptors for pathogens including microorganisms, parasites and viruses, blood group antigens are capable of playing a dominant role in infection pathogenesis [11]. In this study, we found that infections were more prevalent in patients with blood group B (+) ve and O (+) ve; whereas patients with blood group A (-) ve and B (-) ve had a lower frequency of infection. B(+) ve and O (+) ve blood group has previously been reported as the most common blood group within the population of Bangladesh [23] which could be a contributing reason for the higher percentage of these two blood groups among the patients of this study. On the other hand, a number of global studies showed that blood group A was associated with a higher risk for acquiring COVID-19 compared with non-A blood groups, whereas blood group O was associated with a lower risk for the infection compared with non-O blood groups [24, 25]. Our study did not find any similar relationship in regards to the aforesaid fact. Further experiments are required to better understand the associations between Rh (D) blood groups, diabetes and COVID-19.

The clinical manifestations of COVID-19 were almost similar in the group with and without diabetes (Table-2). According to the present study, occurrence of asymptomatic infection is less in COVID-19 patients with diabetes. Compromised immune response of cases with diabetes has been reported to play a role in disease expression and progression [26] and that can be a strong determinant for higher frequency of symptomatic COVID-19 cases. However more immunological investigations of the patients are needed to explain this in detail.

Common symptoms in both groups regardless of age and sex were fever, cough, aches and pain, loss of taste and smell, breathing difficulties. A number of studies previously conducted also support this pattern of symptoms for people with diabetes [6, 13]. Prevalence of fever was found in 75% of the cases, the rate being significantly higher in patients with diabetes than in patients without diabetes. Since diabetes results in an individual’s body to go into an immunocompromised state the odds of secondary infection and by extension, persistence of fever are high [11]. Additionally, breathing anomaly was significantly higher in patients with diabetes. Severe progression of COVID-19 (resulting in a severe case of pneumonia, ARDS) and complex clinical attention such as ICU requirement for individuals diagnosed with COVID-19 are associated with the individual having or not having diabetes mellitus [27, 28]. Excessive uncontrolled inflammatory response, release of tissue injury related enzymes and hypercoagulable state associated with dis-regulated glucose metabolism-all these factors contribute in making an individual diagnosed with COVID-19 having diabetes more susceptible to inflammatory storms which lead to respiratory distress [13].

In terms of comorbidities, cases with diabetes have a higher rate of cardiovascular disease, respiratory disease, cancer and other chronic illnesses like CKD. Microvascular and macrovascular complications of diabetes result in cardiac, renal and neurological diseases. Previous studies have reported similar complications among the same cohort in other countries [29, 30]. Suffice to say, the depletory impact of COVID-19 is higher in comorbid patients than those who do not have comorbidities [31].

Biochemical diagnostic assays reported that significantly greater hyperglycemia was observed in patients with diabetes than those without (Table 2). In cases with diabetes mellitus we found higher concentrations of leukocyte, c-reactive protein and lower lymphocyte percentages compared with cases of COVID-19 without diabetes. Apart from the upper respiratory epithelium & alveolar epithelial cells in lungs, SARS CoV-2 also infects and localizes in immune cells (CD3, CD4, CD8 T cells) inducing lymphocyte apoptosis, suppress innate immunity and enhance cytokine storm [11]. Degree of Lymphocytopenia is associated with the severity of SARS-CoV 2 infection [32-34]. Neutrophil chemotaxis, phagocytosis and intracellular microbe demolition is inhibited by DM. Patients with DM have blunted antiviral INF response delayed activation of Th4/Th17 may contribute to accentuated inflammatory response as is seen in this study by increasing CRP. CRP and ESR is significantly increased among patients with diabetes than those without [6, 13]. High CRP on admission among diabetic patients is a strong indication for severe disease [6] might be explained by the fact that patients with diabetes were more susceptible to pathogens post viral infection due to depleted levels of immune function. Higher levels of D-dimer on admission were found in individuals with diabetes than those without. Similar patterns were also observed by Guozhen et al. (2020) in their study [35]. COVID-19 patients with diabetes had elevated levels of D-dimer, an activation marker of fibrinolysis which is a significant prognostic factor in patients of pneumonia sepsis [36, 37]. In this study, compared to COVID-19 patients without diabetes, geriatric patients with diabetes had significantly higher levels of D-dimer. This factor might alter immunological & pulmonary functions in individuals with diabetes. To better understand the mechanism, further extensive studies and analyses are required.

Compared with COVID-19 patients without diabetes, Troponin-1 was also found to be present at significantly higher levels in those with diabetes (Table 3). Findings similar to ours were found in Italy previously [38]. Considerable proportion of patients presented with elevated cardiac troponin level in patients with comorbidities including diabetes mellitus [38]. Identification of high mortality risk in older adults might be possible by high sensitivity cardiac troponin and it might also be useful when it comes to scrutinizing the clinical fundamentals of older adults with diabetes. However, this retrospective study had a smaller size in terms of cases with diabetes therefore it may lead to a selection bias. Thus, further studies are required for a better perception.

Besides aggravating pre-existing diabetes, COVID-19 can also trigger new onset of Diabetes Mellitus. In this study, we found a few numbers of new cases of diabetes (1.34%, n=10) post COVID-19 infection. Although the presence of ACE2 receptors were found on the pancreatic beta cells [39], however, whether it was Type 1 Diabetes Mellitus or Type 2 Mellitus could not be shed light upon and thus calls for further analyses and cohorts to assess.

The frequency of patients taking insulin during COVID-19 increased more than three-fold in terms of the intake frequency prior to COVID-19 occurrence (Figure 1). Angiotensin converting enzyme 2 (ACE 2) has a potentially important molecular link between insulin resistance and COVID-19 severity during COVID-19 induced cytokine storm where increased insulin resistance induces tremendous insulin requirement [40]. In patients with DM, any infection can worsen glycemic control through stress, mediated through mechanisms such as enhancement of cortisol of release. This would be exacerbated by any uses of exogenous glucocorticoid therapies [41]. In this study about one quarter of patients took steroids. Use of steroids during COVID-19 infection further deteriorates glycemic status as well raising insulin requirement. Majority of patients get > 40-unit insulin during COVID-19 i.e. 40 unit/Ks/ day. Similar results were also found in other studies [42]. Intensive insulin treatments, such as basal bolus insulin therapy and continuous intravenous insulin infusion are known to be out of harm’s way and effective therapeutic measures when it comes to the management of hospitalized patients having hyperglycemia [43, 44].

Metformin is the basal therapy for Type 2 diabetes mellitus cases often in combination with other therapeutic measures. However, metformin has a potential side effect in lactic acidosis, with heightened risk in the context of renal, cardiac and liver impairment, hypertension and acute illness. Therefore, current NICE guidelines recommend temporary discontinuation of metformin therapy during acute illness (including COVID-19 infection) [45]. But in this study, about half of the patients continued taking metformin while they were diagnosed with COVID-19 as well as sulfonylurea. Though DPP4 inhibitor is preferably safe, its frequency of intake is reduced in this study in individuals with COVID-19. Among the other drugs that included SGLT2 inhibitors the intake was reduced which is beneficial for patients with DM & COVID-19. SGLT2 inhibitors may cause dehydration as well as increase risk of Euglycemic ketoacidosis [46].

Regarding usage of antibiotics the majority of patients took azithromycin either by their own self of judgement or after consulting with a physician. On the other hand, few patients took Doxycycline and Ivermectin despite lack of clinical evidence supporting their therapeutic effectiveness against COVID-19. Most recent guidelines suggest avoiding use of antibiotic agents blindly and should only be used when there are confirmed secondary bacterial infections [47]. However, when bacterial infections are identified, rational use of antibiotic agents showed valid results in relieving symptoms & reducing leukocyte count [48].

To our knowledge, this is the first systematic cohort study of the psychiatric and physical consequences of coronavirus infection among Bangladeshi COVID-19 patients with diabetes. Some recent studies reported that the common physical symptoms of COVID-19 patients are fatigue, dyspnea, joint pain, and chest pain [14]. We have observed that complexities such as mobility impairment and pain were prominent in COVID-19 patients with diabetes. A number of evidence linked vitamin D deficiency to COVID -19 morbidity and mortality [19, 49]. Multiple organ damage induced by COVID-19 might be prevented by vitamin D supplementation [19]. Chronic pain comes with psychological distress accompanied with physical disability among its symptomatic manifestations which can be exasperating for patients with diabetes [4]. Our main findings are similar to previous observations in the acute stage of SARS, MERS, and COVID-19 [50]; there is evidence of depression, anxiety, fatigue, and post-traumatic stress disorder in the post-illness stage of previous coronavirus epidemics [5]. Our observations on COVID-19 diabetic patients are in line with these previous observations. Other common psychiatric findings were depression (24%), panic attack (13%), loss of concentration (29%), loss of memory (22%), and insomnia (13%). Notably, insomnia, memory loss, anxiety, and depression were relatively common, suggesting that although a full syndrome of mania was uncommon, subthreshold symptoms might be present. The frequency of anxiety, depression, and post-traumatic stress disorder were high, although the lack of adequate comparison groups or assessment of previous psychiatric disorder was absent in this study. Thus, it is hard to separate the effects of the infection from the impact of an epidemic on the population as a whole or the possibility that selection bias led to the high prevalence figures. In cohorts with SARS, measures of health-related quality of life were considerably lower as compared to those in control groups. However, effects on social functioning were more impaired as compared to that on mental health. However, the common symptoms that were reported to be highly prevalent (such as depression, anxiety, loss of concentration, memory loss and fatigue) could implore selection bias and be irrelevant to SARS-CoV-2 infection. Since our study results were based solely on cohort information, it was not confirmed by any professionals. So further psychiatric, clinical and molecular research is required to explain the mechanism of various abnormalities due to COVID-19.

## 5.0 Limitations

The study population only included patients from four selected COVID-19 hospitals in Bangladesh and no community population was enrolled in the research study. There could be possibilities of information bias as well as recall and interviewer bias because the interviews were conducted over the telephone. Data related to biochemical assays were not available for more than 50% of the patients. During the early stages of the pandemic, diagnostic facilities for COVID-19 cases were scarce, so were the financial means for some individuals which held them back from going for any kind of biochemical tests and evaluations. Regardless, this study is the most substantial one conducted from Bangladesh on COVID-19 to date.

## 6.0 Conclusion

COVID-19 patients with diabetes were more likely to be symptomatic and within higher age groups. Cohorts with diabetes exhibited and nested more comorbidities and biochemical aberrations than cases without diabetes. A number of long-term complications were observed among recovered COVID-19 patients with diabetes. Further molecular, psychological and clinical studies are required to better understand the relationship between COVID-19 and diabetes.

## Data Availability

Data is available upon request from corresponding author.

## 7.0 Acknowledgement

Authors would like to thank the volunteers of Disease Biology and Molecular Epidemiology (dBme) Research Group, Chittagong for their support to conduct the study.

